# Abnormal temporal prediction relates to psychomotor retardation in major depressive disorder

**DOI:** 10.1101/2025.07.08.25331088

**Authors:** Jianfeng Zhang, Yingying Wang, Jingyu Hua, Yuqi Zhang, Jing Yuan, Jinfang Han, Zhonglin Tan, Nai Ding, Georg Northoff

**Author notes:** Corresponding addressed to: Nai Ding, Key Laboratory for Biomedical Engineering of Ministry of Education, College of Biomedical Engineering and Instrument Sciences, Zhejiang University, Hangzhou, China, Zhonglin Tan, Affiliate mental health center and Hangzhou Seventh People’s Hospital, Zhejiang University School of Medicine, Hangzhou, China.

## Abstract

Major depressive disorder (MDD) exhibits psychomotor retardation which concerns abnormal slowness of both thoughts and movements. In the current study, we investigated whether such psychomotor disturbance also disrupts the MDD subjects’ ability to synchronize their movements to a rhythmic tone sequence. We found that the timing between finger taps and tones is altered for MDD subjects (N=38), compared with healthy controls (N=65), mainly when tones are presented at a low frequency below 1 Hz. No differences, however, are observed in terms of the tapping speed, tapping stability, and tapping interval. Auditory-motor desynchronization in the low frequency range (< 1Hz) is associated with depression severity as well as with the degrees of both psychomotor anhedonia and vegetative symptoms (using the Beck Depression Inventory). In sum, our study demonstrates a frequency-specific auditory-motor synchronization deficit in MDD which also relates to psychomotor retardation. This suggests disruption of the cognitive capacity of temporal prediction for low-frequency auditory-motor synchronization in MDD. That supports a cognitive and therefore psychomotor over a motor view of psychomotor retardation in MDD.

## Introduction

Psychomotor retardation, characterized by a slowing of thoughts, speech, and body movements, is a core symptom of major depressive disorder (MDD) (1–5). Traditionally assessed through subjective depression rating scales (1, 3–7) or objective measures like reaction time tasks, actigraphy, and speech analysis (1, 8–11). Recent research suggests that psychomotor retardation may have broader cognitive implications beyond overt motor function, based on the findings of the complex interaction between dopamine-based subcortical-cortical motor circuits (1, 2, 9–15) and other neural networks (2, 12, 13), which points to a more intricate relationship between motor and cognitive processes in MDD. However, the specific mechanisms by which motor circuit dysfunction influences non-motor cognitive functions remain poorly understood, highlighting the need to investigate the cognitive contributions of motor system to non-motor function in both healthy individuals and those with MDD.

Recent evidence suggests that the motor system may function as a rhythmic generator, providing temporal prediction, particularly for the auditory modality (16–20). Neurobiologically, the motor system is coupled with the auditory neural pathway (19, 21, 22), as evidenced by the recruitment of motor system during passive listening tasks, even when attention is directed away from auditory stimuli (23, 24). This reveals an automatic process wherein motor regions track temporal features in auditory streams (19). Motor system do contribute to auditory perception in a frequency-specific manner, particularly facilitating the processing of low-frequency information through temporal prediction (16, 17, 21, 25). Specifically, motor regions transmit signals to auditory areas, predictively aligning ongoing low-frequency oscillations with anticipated events, thereby enhancing auditory perception (16, 17). In short, the motor system provides temporal predictions to sensory regions like the auditory cortex to optimize its processing at specific frequencies (16). While this is well known in the healthy brain, how such auditory-motor temporal prediction is altered in MDD and especially its relationship to psychomotor retardation remains yet unclear though.

One behavior measurement related to the temporal prediction is auditory-motor synchronization (AMS) (26, 27). AMS refers to the coordination of rhythmic body movements with an external auditory rhythm, exemplified by tapping fingers to the beat of music (26–30). This process relies on the ability to perceive beats in auditory signals, generate internal temporal predictions or expectations, and utilize feedback to plan synchronized motor responses, thus engaging distributed brain networks (31). Neuroimaging studies have delineated the neural basis of AMS. Simple synchronized tapping activates motor areas including the cerebellum, basal ganglia, premotor and supplementary motor cortices, thalamus, and auditory regions (27, 30, 32). More complex AMS tasks engage non-primary motor areas to a greater extent than self-paced movements, reflecting increased demands on timing, sensorimotor integration, and audio-motor coupling (33, 34). While previous studies using finger-tapping tasks have yielded non-significant results (35–37), they did not test the frequency specificity for abnormal temporal prediction in auditory-motor synchronization. Addressing this yet open question is the objective of our study.

In this study, we hypothesize that AMS abnormalities may exist in MDD but in a frequency-specific manner, given that psychomotor retardation is a key deficit in MDD and is related to altered neural activity in motor system (29). We aim to investigate whether the performance of auditory-motor synchronization can reveal abnormal temporal prediction in MDD and whether it relates to psychomotor retardation with frequency-specific patterns. To this end, we employed a standard click-tapping paradigm (26). Participants were instructed to tap in synchrony with a tone sequence, with rhythmic frequencies ranging from 160 ms to 2600 ms in inter-onset interval (IOI) across different tone sequence trials (Figure 1). Psychiatric scales were also administered. Performance was analyzed to determine whether MDD show altered auditory-motor synchronization performance, and if so, whether these alterations occur in a frequency-specific manner. Importantly, we conducted extensive control analyses to account for attention/motivation and executive capacities in order to sharpen the link of the assumed potential deficit in auditory-motor synchronization with altered temporal prediction. Finally, we examined how alterations in auditory-motor synchronization correlate with psychomotor retardation. In sum, our study not only seeks to quantify the extent of psychomotor retardation in a frequency-dependent manner but also to understand how alterations in AMS relate to the broader symptomatology of depression, potentially offering new avenues for targeted interventions.

**Figure 1.**
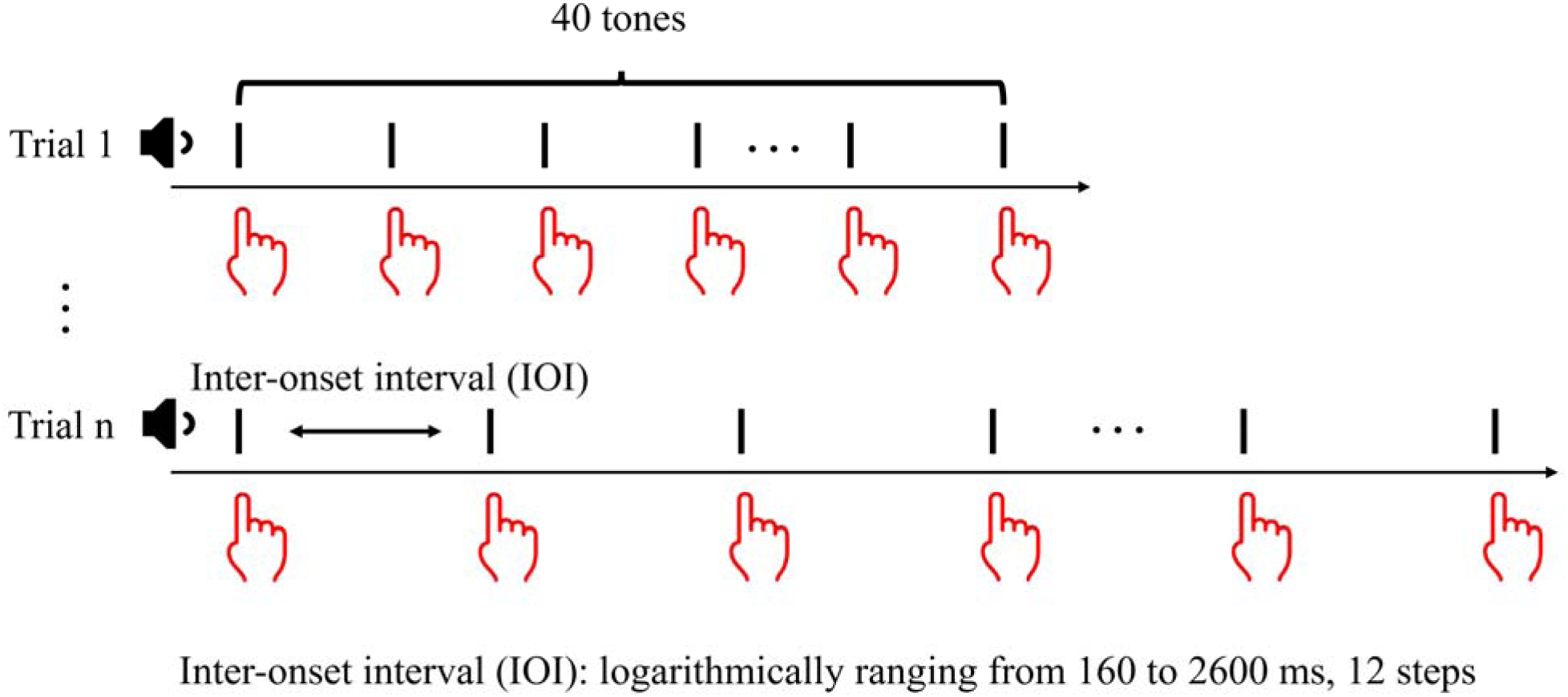
Schematic representation of the auditory-motor click-tap paradigm. Participants were instructed to synchronize their finger taps with a sequence of rhythmic tones. The inter-onset intervals (IOIs) were logarithmically distributed across 12 steps, ranging from 160 ms to 2600 ms. Each IOI condition consisted of three trials, with each trial comprising a sequence of 40 tones (i.e., 120 tones for each condition). Trial presentation order was randomized across conditions.

## Materials and Methods

### Participants and Clinical Assessment

Participants with Major Depressive Disorder (MDD) were recruited from both inpatient and outpatient services at Hangzhou Seventh People’s Hospital, Zhejiang, China. Inclusion criteria specified: (1) age 18-65 years; (2) diagnosis of MDD according to DSM-5 criteria; (3) confirmation of no comorbid mental disorders via Mini International Neuropsychiatric Interview (M.I.N.I.) (38); (4) Hamilton Depression Scale (HAMD) score ≥18; (5) psychotropic medication-free or stable regimen for minimum one week; (6) absence of benzodiazepine use; (7) stable mood stabilizer dosage for minimum two weeks; and (8) normal thyroid function. Healthy controls were recruited according to the following criteria: age 18-65 years, absence of self-reported psychiatric disorders (including psychotic, mood, or anxiety disorders), no substance abuse history.

The final sample comprised 38 MDD patients (36 female, age range: 18-34 years) and 65 healthy controls (61 female, age range: 18-26 years). While groups were matched for gender distribution, a significant age difference was observed between groups (p<0.01). Musical training experience was documented due to its potential influence on click-tapping performance. The Subject demographic and clinical information can be observed in Table 1.

**Table 1:** Subject demographic and clinical information.

| Variables | MDD (n=38) | HC (n=65) | Statistics | p-value |
| --- | --- | --- | --- | --- |
| Age (years) | 22.03 ± 3.40 | 20.60 ± 1.63 | t = 2.87 | 0.005 |
| Sex (Female/Male) | 36/2 | 61/4 | $\chi^2 = 0.05$ | 0.823 |
| Music Training (years) | 0.28 ± 1.09 | 0.24 ± 0.76 | t = 0.21 | 0.837 |
| HAMD-17 Score | 22.34 ± 3.69 | - | - | - |
| BDI Total Score | 30.79 ± 8.05 | 7.14 ± 5.56 | t = 17.59 | <0.001 |

Data are presented as mean ± SD. MDD = Major Depressive Disorder, HC = Healthy Controls, HAMD-17 = 17-item Hamilton Depression Rating Scale, BDI = Beck Depression Inventory

### Ethical statement

The authors assert that all procedures contributing to this work comply with the ethical standards of the relevant national and institutional committees on human experimentation and with the Helsinki Declaration of 1975, as revised in 2013.

All participants provided written informed consent prior to study enrollment. All procedures have been approved by the Institutional Review Board, and comply with the ethical standards of the Hangzhou Seventh People’s Hospital.

### Paradigm design

The study employed an auditory-motor click-tap paradigm (Figure 1). Each trial consisted of 40 tones (50ms duration, 5ms ramp) presented at a comfortable volume. Participants were instructed to synchronize their tapping with the rhythmic tone sequences. Inter-onset intervals (IOIs) were logarithmically distributed across 12 steps ranging from 160ms to 2600ms. Each IOI condition comprised three trials, with trial order randomized across conditions.

### Data analysis

Mismatch asynchrony (MA) was calculated as the temporal difference between each tap and its closest corresponding tone. Taps occurring beyond 50% of the IOI were classified as omissions, and set as NaNs in data analysis. Mean MA was computed by averaging measurements from the 6th to 40th tone, following the rhythm acquisition period (i.e., 1st to 5th tone). To control for potential confounds related to task motivation, particularly during low-frequency conditions, we analyzed MA relative to predicted tones—observable only during maintained rhythmic prediction.

To differentiate between psychomotor function and motor executive function effects on group MA differences, we assessed three parameters: tapping reaction time (initial MA), tapping stability (MA standard deviation), and rhythm asynchrony (interval disparity between taps and tones) (see Figure 3A).

### Beck depression inventory (BDI) and its subscales assessment

The Beck Depression Inventory-II (BDI-II) was administered to assess the presence and severity of depressive symptomatology (Beck et al., 1996). The BDI-II comprises 21 self-report items, each rated on a four-point ordinal scale (0–3), where "0" indicates absence of symptoms (or "as usual" functioning), and scores of 1-3 represent increasing symptom severity. The instrument demonstrates robust psychometric properties, with items exhibiting strong reliability and appropriate distribution across the depressive symptom continuum(39).

Factor analytic studies (40, 41) have established a four-dimensional structure of the BDI-II (i.e., four subscales). The first dimension, negative cognition, encompasses six of the eight cognitive symptoms: past failure (#3), guilty feelings (#5), punishment feelings (#6), self-dislike (#7), self-criticalness (#8), and worthlessness (#14). The remaining three dimensions reflect somatic-affective components, they are: psychomotor anhedonia: comprising anhedonia (#4), irritation (#11), social anhedonia (#12), difficulty in decision-making (#13), and reduced work capacity (#15); vegetative symptoms: encompassing late insomnia (#16) and appetite loss (#18); and somatic symptoms: including weight loss (#19) and health-related concerns (#20). This dimensional structure provides a comprehensive framework for assessing both cognitive and somatic-affective manifestations of depression, enabling more nuanced analysis of symptom patterns and severity.

### Statistical analysis

All analyses were conducted using MATLAB. Group comparisons of MA and executive function were performed using two-sample t-tests. To address potential confounding effects of age differences between groups, we implemented linear mixed-effects models (LME) using the fitlme function from MATLAB’s Statistics and Machine Learning Toolbox. The primary model incorporated fixed effects (Group, IOI, and Group×IOI interaction), subject-level random effects, and age as a covariate: MA ∼ Group*IOI + Age + (1|SubjectID). To evaluate the contribution of age to the model, we constructed a reduced model without the age variable: MA ∼ Group*IOI + (1|SubjectID), and compared these nested models using likelihood ratio tests.

Chi-square analysis was employed to assess group differences in missing tone response rates. Correlations between MA and BDI scores (including subscales) were evaluated using Pearson correlation coefficients. Multiple comparisons were controlled using False Discovery Rate (FDR) correction within each analysis.

## Results

### Altered auditory-motor synchronization during slow speed

To investigate the group differences in auditory-motor synchronization during different timescales, we adopted a standard click-tapping paradigm (26, 27). The subjects were asked to tap in synchronization with the rhythm of tone sequences. Each tone sequence contained 40 tones in total whose rhythmic frequency ranged from 160 ms to 2600 ms in inter-onset interval (IOI) during different tone sequence trials (Figure 1).

The performance of auditory-motor synchronization was investigated by the tapping performance in relation to the isochronous acoustic stimulus, i.e., the asynchrony between tapping and tone. Once the subjects can entrain to the isochronous auditory stimuli, they tend to anticipate the next tone with their motor tap preceding the timing of the subsequent auditory input (26, 27). After removing the first five tones (as needed for entraining subjects to auditor-motor synchronization), we observed that the MDD patients (N=38) and HC (N=65) exhibited similar mean asynchrony (MA) at the fast frequencies. The mean asynchrony in both turned to negative at the speed of 339 ms in the IOI. The pattern of group differences demonstrated frequency specificity, with the majority of fast conditions (i.e., IOI < 1000 ms) showing no significant alterations, although one exception was observed at IOI = 339 ms. In contrast, systematic and robust group differences consistently emerged across the slow conditions (i.e., IOI > 1000 ms). Replicating previous findings of sensory-motor asynchrony (27) (42), healthy subjects showed negative asynchrony in the low frequencies (< 1Hz); this means that they tended to tap early relative to the external auditory tone with the tap preceding the latter, a phenomenon indicative of temporal prediction. In contrast, MDD showed decreased negative asynchrony (i.e. tapping occurring close to the tone onset rather than preceding it) in the same low frequencies (Figure 2B); this means that, unlike healthy subjects, MDD subjects did not tend to tap early preceding the tone – this is indicative of a deficit in temporal prediction in especially the lower frequencies.

**Figure 2.**
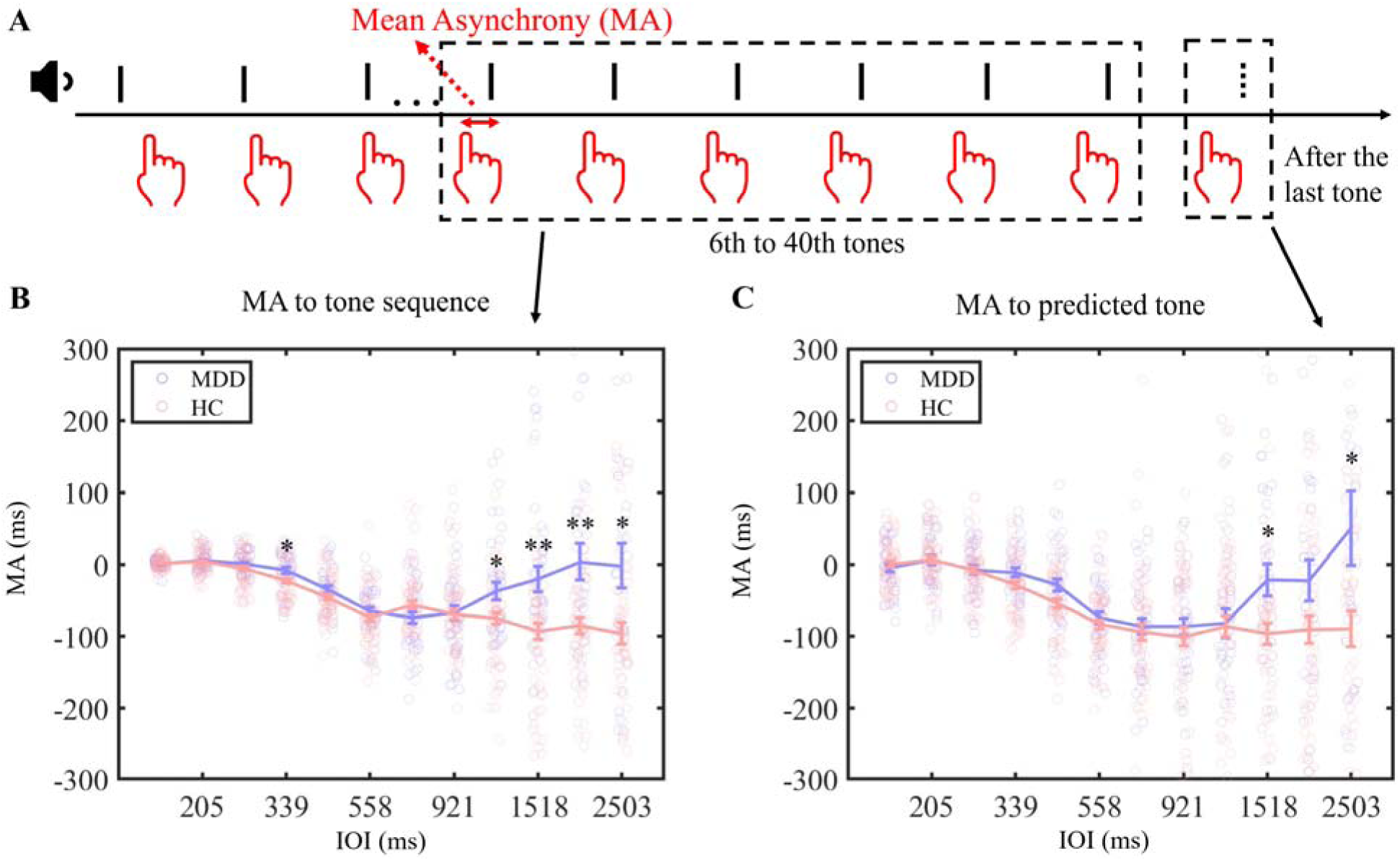
Comparison of Mean Asynchrony (MA) Between Healthy Controls (HC) and Major Depressive Disorder (MDD) Participants Across Temporal Frequencies. (A) Methodological schematic of MA calculation. MA was defined as the temporal interval between each tap and its corresponding tone. Initial synchronization period (first 5 tones) was excluded from analysis. MA for tone sequences represents the average asynchrony from tones 6-40. MA to predicted tone was calculated as the temporal deviation from the expected (virtual) tone following the sequence termination. (B) MA analysis for tone sequences. Significant group differences emerged exclusively in the low frequency range (IOI >1000 ms). Error bars represent standard error of the mean. Statistical comparisons were conducted using two-sample t-tests for each IOI, with p-values adjusted for multiple comparisons using FDR correction. (C) MA analysis for predicted tones. Results parallel the pattern observed in tone sequences, with group differences predominantly in the low frequency range. Error bars represent standard error of the mean. Statistical significance was determined using two-sample t-tests for each IOI, with FDR correction for multiple comparisons. *p<0.05, **p<0.01.

### Altered auditory-motor synchronization cannot be explained by lack of attention or motivation

As the mean asynchrony at the low frequencies in MDD was close or even larger than 0, this may reflect a deficit in slow-range audio-motor synchronization, or alternatively, a difference in motivation in conducting the task. For example, in slow conditions, the patients may merely tap along their perception of the tones rather than properly entraining to the rhythm of the tones. To distinguish these two hypotheses, we further investigated the tapping performance with regard to the predicted tone (Figure 2A). The patients failed in higher numbers in response to the predicted tones (in two slowest conditions: IOI = 1949 ms, χ^2^ = 10.9, p < 0.001; IOI = 2503 ms, χ^2^ = 5.3, p < 0.05). This may be related to decreased attention or motivation in which case one would expect a higher number of misses or non-responses in MDD, when the task difficulty increases in the slow frequency.

However, the higher number of failures to response to the predicted tone cannot fully account the group difference of MA to the predicted tone. To control for such potential attention or motivation deficits, we removed the non-responses from our data. This did not affect the main result though, as the MDD subjects’ degree of asynchrony was still significantly lower than in healthy subjects in the low frequencies (p < 0.05, FDR-corrected). This suggests that the decreased performance in asynchrony is mainly related to the deficit in low-frequency entrainment rather than the level of attention or motivation for conducting the task (Figure 2C).

### Altered auditory-motor synchronization cannot be explained by lack of executive capacity

We next investigated whether the altered auditory-motor asynchrony is related to the executive capacity of motor function as measured by reaction time, tapping stability and tapping frequency range.

The reaction time to the auditory input stimuli was measured by the finger tapping in response to the first tone (i.e., initial MA), as here there was not yet any temporal prediction involved. By comparing the reaction time between groups, no difference was observed across all frequencies (including both slow and fast); this suggests intact auditory-motor response function in MDD (Figure 3B).

**Figure 3.**
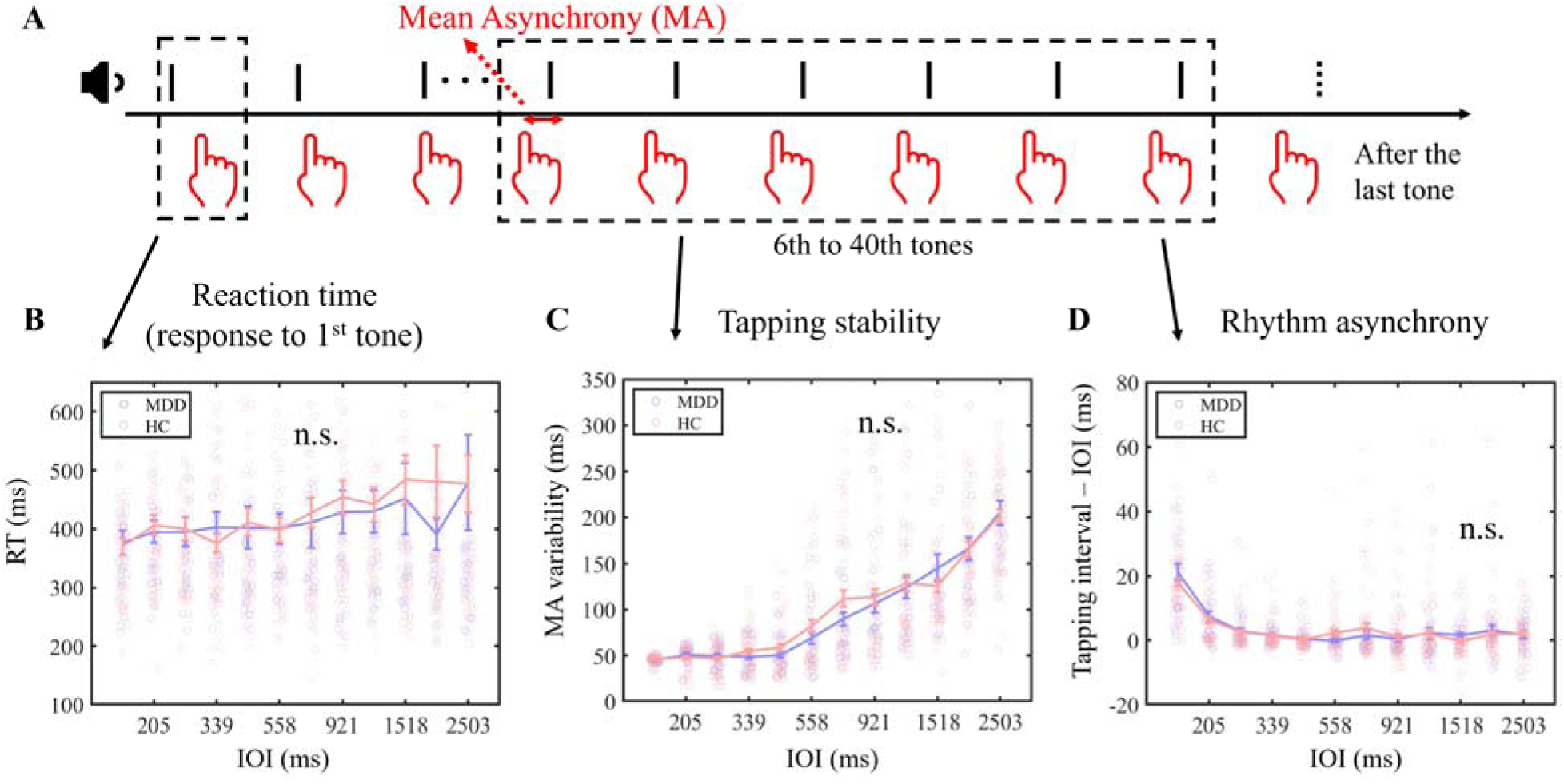
Comparison of Motor Executive Functions Between Healthy Controls (HC) and Major Depressive Disorder (MDD) Participants in the Auditory-Motor Synchronization Task. (A) Schematic representation of three motor executive function parameters: reaction time, tapping stability, and rhythm asynchrony. (B) Analysis of reaction time, quantified by the mean asynchrony (MA) to the first tone, before rhythm synchronization was established. No significant differences were observed between HC and MDD groups across all inter-onset intervals (IOIs). (C) Assessment of tapping stability, measured by the standard deviation of MA from tones 6-40. No significant group differences were detected across the IOI range. (D) Evaluation of rhythm asynchrony, calculated as the temporal interval difference between tapping and tone sequences from tones 6-40. Analysis revealed no significant differences between groups across all IOIs. Statistical comparisons were conducted using two-sample t-tests for each IOI, with p-values adjusted for multiple comparisons using FDR correction. Error bars represent standard error of the mean.

Secondly, we investigated the tapping stability by measuring the standard deviation (SD) of the subjects’ asynchrony time in their tapping. SD as proxy of tapping stability increased with prolonged IOI in both MDD and healthy subjects, but no significant difference between groups was observed. This suggests that MDD do not suffer from a deficit in their tapping stability during both slow and fast frequencies (Figure 3C).

Lastly, we investigated the capability of synchronization range by measuring the difference of IOI between the input (i.e., tone sequence) and the output (i.e., tapping). Both groups failed to entrain to the fastest conditions (i.e., IOI: 160 ms and 205 ms, p < 0.01 in both, FDR-corrected). However, no group difference between MDD and HC was observed. This suggests that MDD subjects do not suffer from an input processing deficit (Figure 3D).

### Effects of age on auditory-motor synchronization

As illustrated in Table 1, we observed that the two groups differed in ages. To evaluate whether age differences between groups might account for the observed auditory-motor synchronization patterns, we conducted linear mixed-effects model analyses. Comparison of models with and without age as a covariate revealed that including age did not significantly improve model fit (LRT=3.1318, df=1, p=0.0768), indicating that age differences did not substantially influence auditory-motor synchronization performance when accounting for within-subject variability.

### Altered auditory-motor synchronization relates to psychomotor retardation

Lastly, we investigated whether the altered mean asynchrony was related to clinical symptoms like psychomotor retardation in MDD. We first correlated the mean asynchrony with the total score of the Beck depression inventory (BDI); we observed that only the asynchrony performances at the low frequencies (< 1Hz) (but not the ones at faster frequencies) correlated with the total BDI score. This was followed up by investigating the correlation with the BDI subscales. We observed that psychomotor anhedonia and vegetative symptoms correlated with the mean asynchrony at the low frequencies (< 1Hz) (Figure 4), suggesting that the AMS deficit mainly relates to those depressive symptoms that are associated with psychomotor and bodily dysfunction. The scatter plot of these correlations can be observed in supplementary materials (Fig. s1 - Fig. s5).

**Figure 4.**
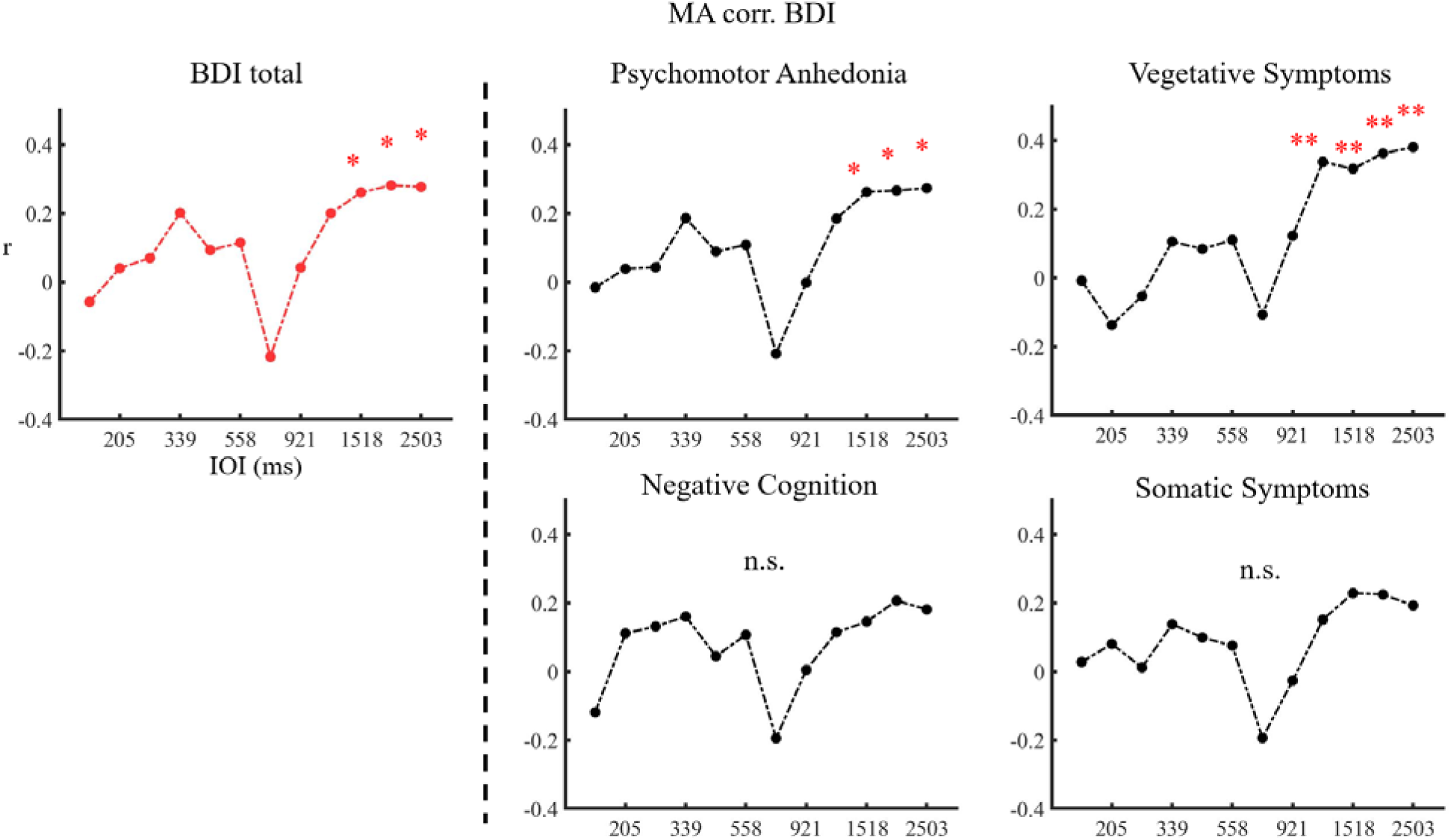
Associations Between Mean Asynchrony (MA) and Beck Depression Inventory-II (BDI-II) Dimensions in the Auditory-Motor Synchronization Task. Analysis of correlations between MA and BDI-II total scores and its four constituent dimensions: psychomotor anhedonia, vegetative symptoms, negative cognition, and somatic symptoms. Correlation analyses were performed across all inter-onset intervals (IOIs) at the individual participant level. Statistical significance was determined using Pearson correlations with FDR correction for multiple comparisons. *p<0.05, **p<0.01

## Discussion

Our study demonstrates a frequency-specific auditory-motor synchronization deficit in MDD, mainly evident at slower frequencies (<1Hz). This impairment cannot be attributed to deficits in attention, motivation, or executive motor function. Importantly, the observed desynchronization at low frequencies correlates significantly with depression severity, specifically with psychomotor anhedonia and vegetative symptoms. These findings suggest that psychomotor dysfunction in MDD extends beyond purely motor-related symptoms, potentially disrupting the temporal predictive capability from the motor system to other cognitive domains. Our results provide new insights into the nature of psychomotor retardation in depression in that it shows an underlying temporal deficit, that is, temporal prediction on the behavioral level.

### Auditory-motor coupling, the timescale, its cognitive function and clinical relevance

The auditory and motor systems exhibit robust coupling across multiple timescales (16, 18, 21), with important implications for cognitive function (17) and clinical applications (43). In healthy individuals, auditory-motor synchronization has been observed across a wide range of frequencies, from very slow rhythms around 0.5 Hz up to faster rates of 6-7 Hz (27). This broad temporal range allows flexible adaptation to different rhythmic contexts, from the slow prosodic patterns in speech to faster syllabic rates. Notably, synchronization appears most stable and precise in the 1-3 Hz range, corresponding to typical speech and music tempos (44). Our findings extend this literature by demonstrating altered auditory-motor coupling specifically at low frequencies below 1 Hz in individuals with major depressive disorder, suggesting a potential disruption of temporal prediction mechanisms operating over longer timescales.

The cognitive functions subserved by auditory-motor coupling are multifaceted. At a basic level, it allows predictive timing of sensory events, optimizing perception by aligning neural excitability with incoming stimuli (16). Beyond this, auditory-motor interactions play a crucial role in speech perception and production, musical performance, and general temporal processing (31). The motor system appears to provide an internal model or temporal template that shapes auditory predictions and attentional focus (18). Our results highlight how this temporal predictive function may be impaired in depression, potentially contributing to difficulties in sustained attention and cognitive processing of temporally extended information.

While considerable research has examined auditory-motor coupling in normative samples, there remains a gap in understanding how these mechanisms may be altered in clinical populations. Prior work on motor timing in depression has largely focused on explicit timing tasks rather than implicit sensorimotor synchronization (45). Our study bridges this gap by directly investigating auditory-motor coupling across frequencies in MDD, revealing a specific deficit in low frequency synchronization. Our finding may explain the previous negative results, which finds that the psychomotor retardation is not sensitive to finger tapping performance at the relative fast rhythm (< 1000 ms in IOI) (1, 35), and suggests that the finger tapping task, especially the slow tapping, might be a sensitive behavioral marker to psychomotor dysfunction. Additionally, examining auditory-motor interactions across other psychiatric and neurological conditions may yield insights into shared or distinct timing-related pathophysiology in the future.

### Temporal prediction and its linkage to psychomotor rather than motor dysfunction

Our study highlights the critical distinction between psychomotor and motor dysfunction in major depressive disorder (MDD), particularly in relation to temporal prediction abilities. The deficit in temporal prediction at low frequencies (<1 Hz) we observed in MDD patients reflects a disruption in the integration of cognitive timing mechanisms with motor execution, rather than purely motor impairments. This aligns with the concept of psychomotor disturbance as a core feature of melancholic depression (46, 47). Our results suggest that temporal prediction may serve as a key cognitive process underlying psychomotor symptoms in depression, bridging the gap between cognitive dysfunction and observable motor behavior.

Our paradigm, which requires participants to synchronize motor responses with auditory stimuli across various frequencies, builds upon previous work on time perception in depression(48, 49). The specific deficit we observed in low frequency synchronization suggests that depression may particularly impact the ability to predict and prepare for events occurring over longer time scales. This finding resonates with theoretical models proposing that depression involves a fundamental temporal disturbance in the brain’s predictive processes (50) and offers a potential explanation for the subjective experiences of time slowing and difficulty initiating actions in depression (51).

Our results have important implications for understanding the neural basis of psychomotor symptoms in depression. The involvement of temporal prediction processes suggests that psychomotor disturbances may arise from dysfunction in distributed neural networks rather than localized motor system abnormalities. This is consistent with our neuroimaging findings showing altered balance between sensorimotor and default mode networks in MDD patients with psychomotor symptoms (12, 52). Future research should explore interventions targeting temporal prediction abilities to alleviate psychomotor symptoms in depression.

## Data Availability

All data produced in the present study are available upon reasonable request to the authors.

## Acknowledgements and disclosures

This work was supported by Shenzhen-Hong Kong Institute of Brain Science – Shenzhen Fundamental Research Institutions (2023SHIBS0003), National Natural Science Foundation of China 32201129, General Program of Natural Science Foundation of Guangdong Province of China (2025A1515010558), Shenzhen Science and Technology Program (20220811094132001) and the Start-up Research Grant in Shenzhen University to JZ. And the European Union’s Horizon 2020 Framework Programme for Research and Innovation under the Specific Grant Agreement No. 785907 (Human Brain Project SGA2), the EJLB-Michael Smith Foundation, the Canada Institute of Health Research (CIHR), the Start-up Research Grant in Hangzhou Normal University, and the NFRF, uOBMRI Team grant to GN.

The authors report no biomedical financial interests or potential conflicts of interest.

## Notes

### Competing Interest Statement

The authors have declared no competing interest.

### Funding Statement

This study did not receive any funding

### Author Declarations

Ethical statement The authors assert that all procedures contributing to this work comply with the ethical standards of the relevant national and institutional committees on human experimentation and with the Helsinki Declaration of 1975, as revised in 2013. All participants provided written informed consent prior to study enrollment. All procedures have been approved by the Institutional Review Board, and comply with the ethical standards of the Hangzhou Seventh People's Hospital.

