## supplementary materials for "Abnormal temporal prediction relates to psychomotor retardation in major depressive disorder"

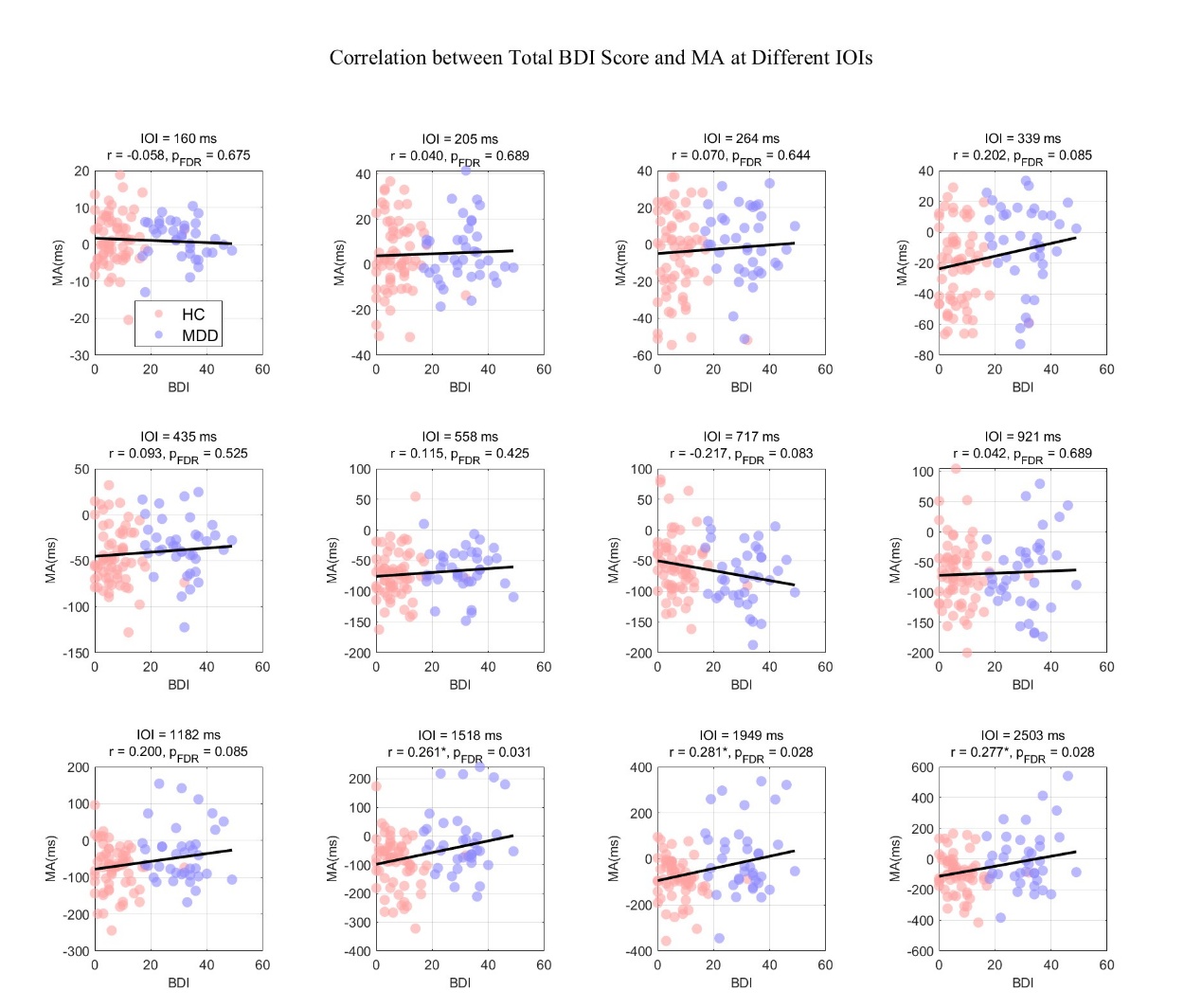

Fig. s1. Scatter plot of correlations between total BDI score and MA at different IOIs.

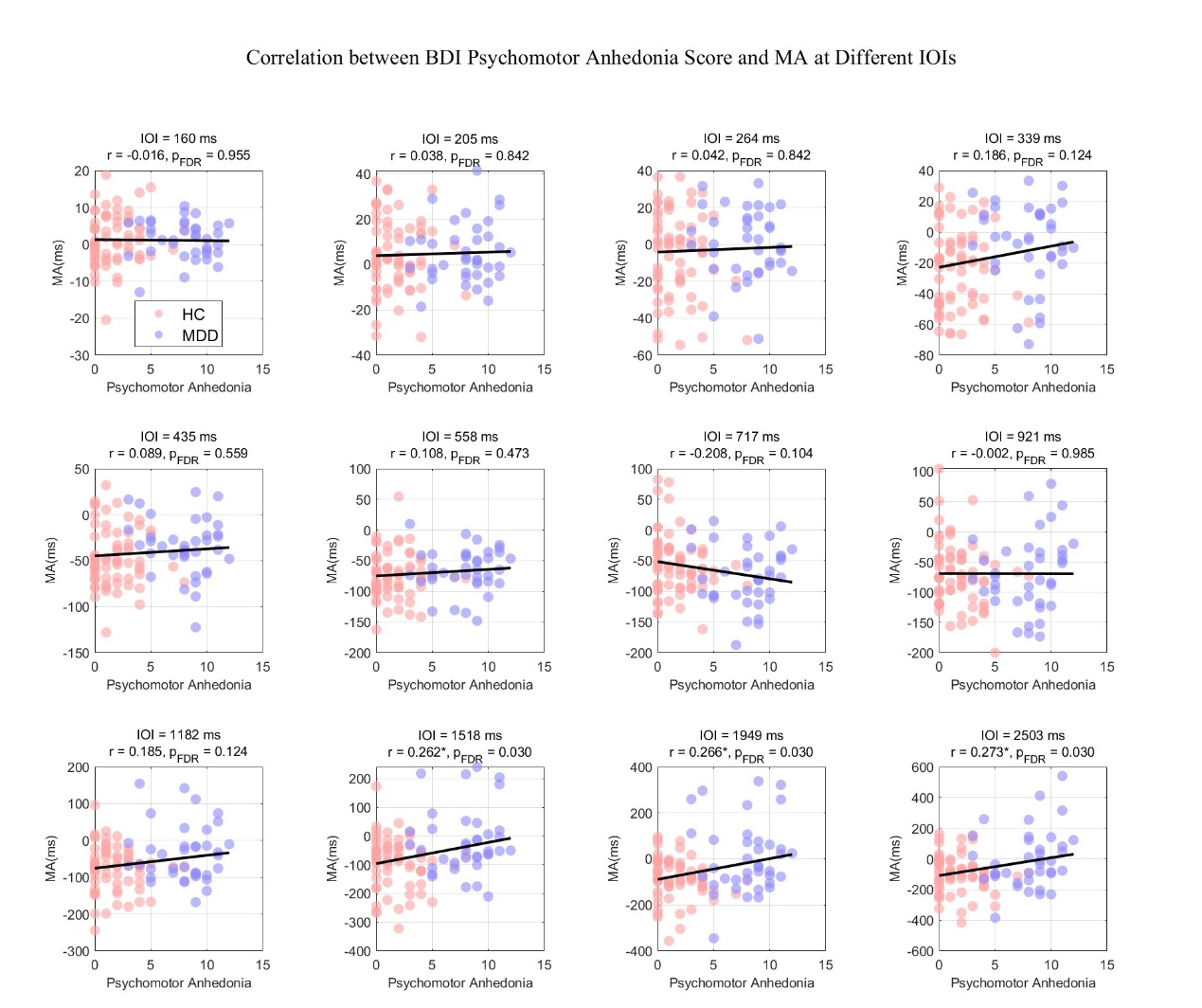

Fig. s2. Scatter plot of correlations between Psychomotor Anhedonia score and MA at different IOIs.

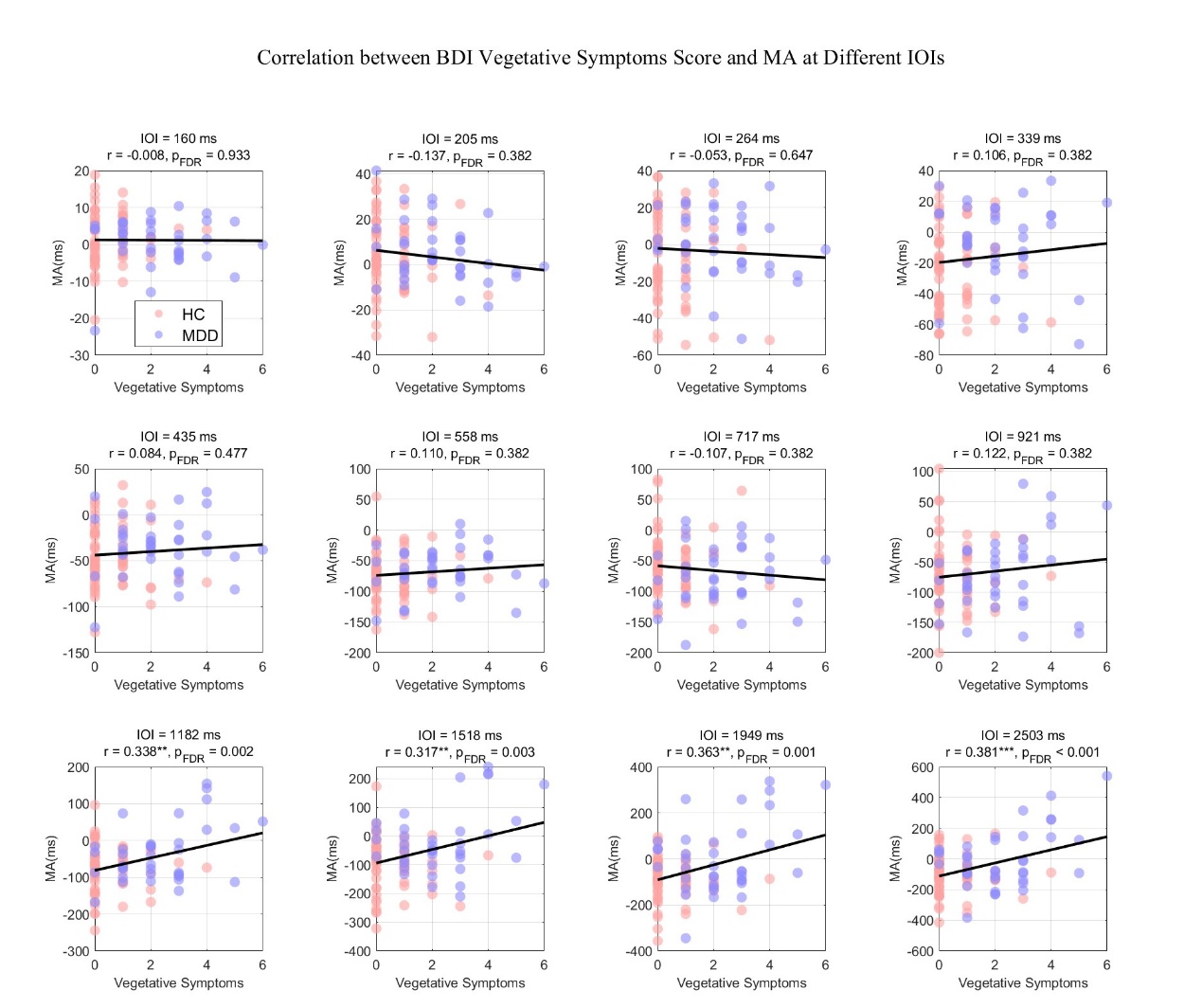

Fig. s3. Scatter plot of correlations between Vegetative Symptoms score and MA at different IOIs.

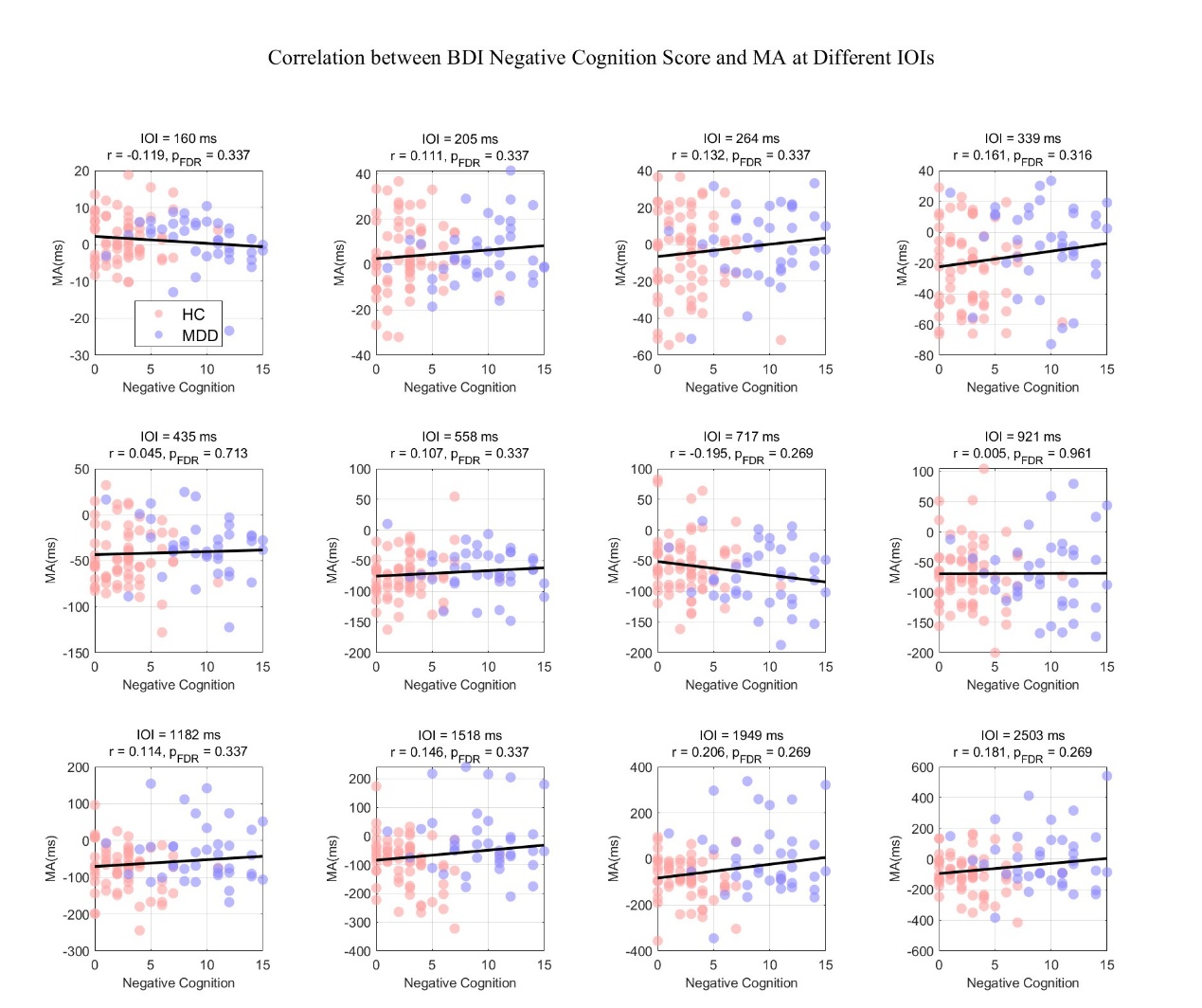

Fig. s4. Scatter plot of correlations between Negative Cognition score and MA at different IOIs.

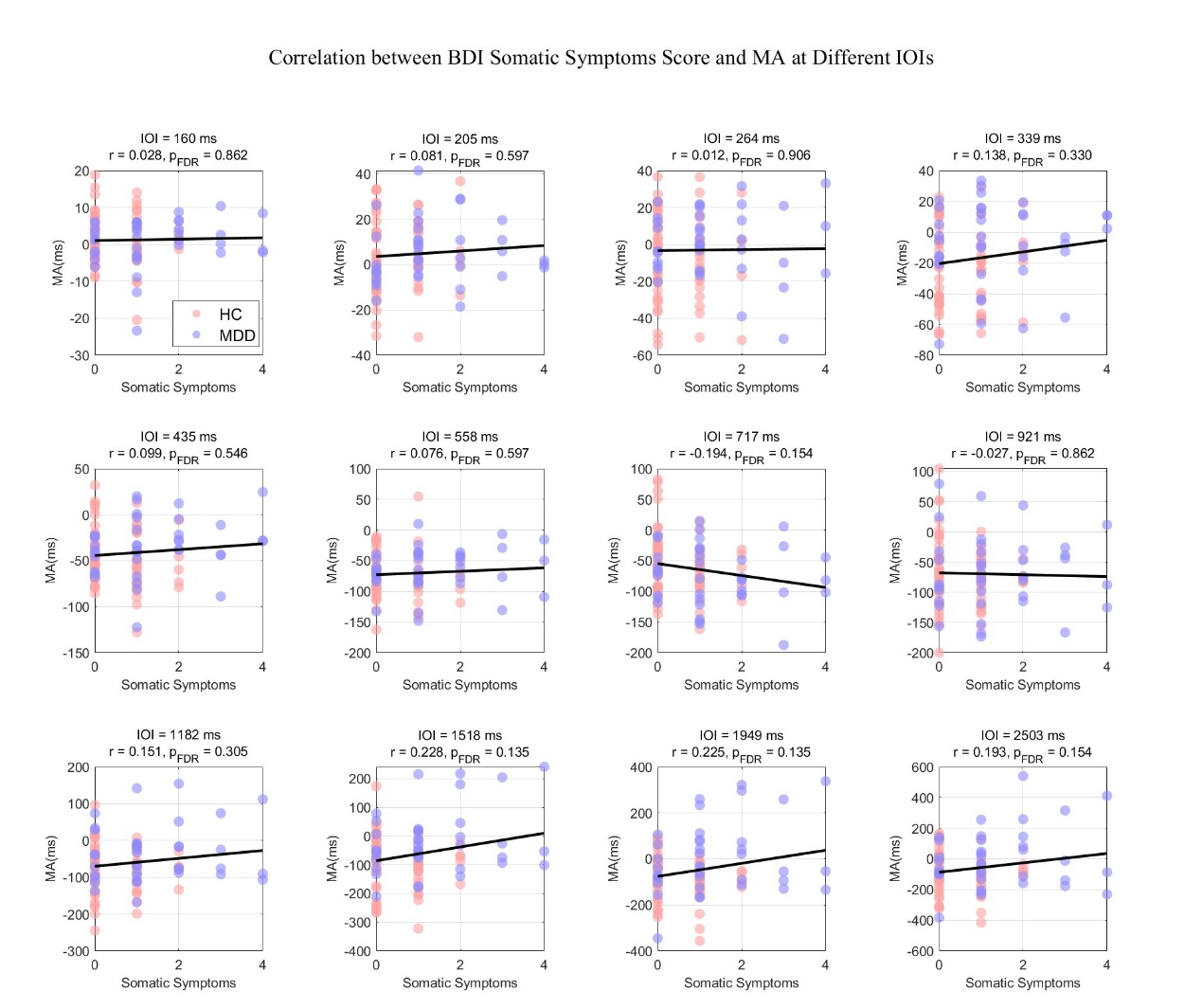

Fig. s5. Scatter plot of correlations between Somatic Symptoms score and MA at different IOIs.
